# EndoGPT: A Proof-of-concept Large Language Model Based Assistant for the Management of Thyroid Nodules

**DOI:** 10.1101/2024.05.29.24308002

**Authors:** Meghal Shah, Eric J. Kuo, Jennifer H. Kuo, Shawn Hsu, Catherine McManus, Rachel Liou, James A. Lee, Tejas S. Sathe

## Abstract

Large language models (LLMs) are increasingly being explored for their potential to simulate clinical reasoning. Here, we demonstrate our initial experience using the GPT-4o LLM along with prompt engineering and knowledge retrieval to develop EndoGPT, a clinical decision support tool for the management of thyroid nodules. In a pilot study of 50 cases, EndoGPT demonstrated an 83% concordance rate with expert surgeons’ assessments and plans. The highest concordance was in diagnosis (93%), followed by the need for an operation (82%) and type of operation (69%). This work suggests that LLM-based assistants may play a useful role in assisting clinicians in the future.

## Introduction

Though large-language models (LLM) demonstrate the ability to answer medical questions, their ability to simulate clinical reasoning is a topic of current exploration. Recent technical advances allow LLMs to be optimized using prompt engineering and knowledge retrieval from data sources, even without specific fine-tuning.^1,2^ Here, we describe our implementation of these techniques to prototype an LLM-based clinical decision support tool for the management of thyroid nodules.

## Methods

We abstracted deidentified data from clinic notes of patients referred for evaluation of thyroid nodules or thyroid cancer. We built an assistant (EndoGPT) based on the GPT-4o LLM that could ingest this data and output a predicted assessment and plan (A&P). To provide EndoGPT with additional context, we uploaded the *2015 American Thyroid Association Management Guidelines for Thyroid Nodules and Differentiated Thyroid Cancer* as a reference.^3^ EndoGPT could then utilize relevant components of the guidelines using vector embeddings and similarity search techniques. ^4^ For each patient scenario, we generated five predicted A&Ps and ensembled them into a compound A&P using a second assistant. After pre-testing EndoGPT on 25 patient scenarios, we analyzed errors, wrote instructions to avoid them, and added this data to EndoGPT’s prompt for additional context before testing it on new scenarios (Figure 1).

**Figure 1:**
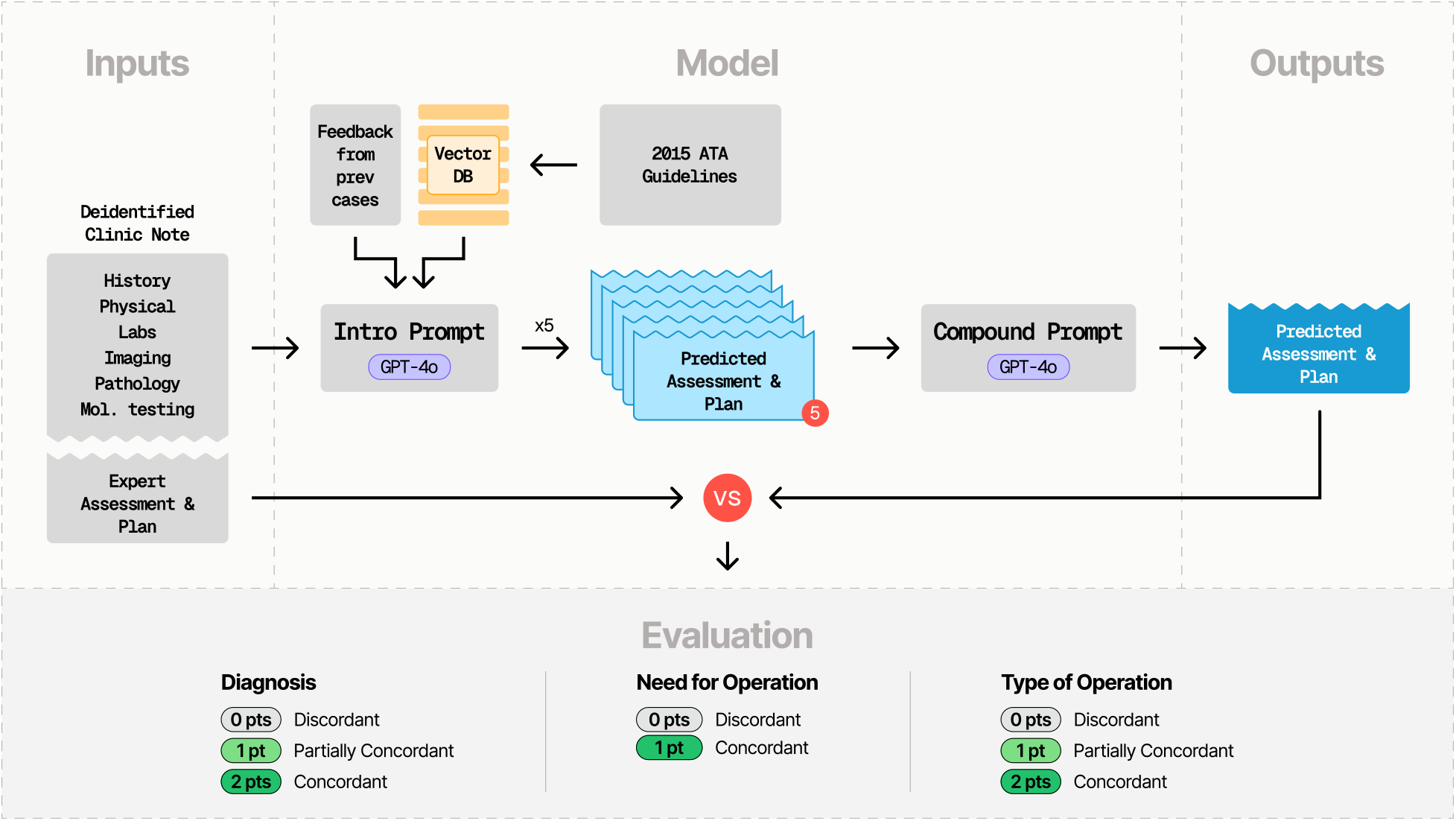
We built an LLM-based assistant called EndoGPT. The input to EndoGPT is a deidentified clinic note excluding the expert surgeon’s assessment and plan. EndoGPT was built using the GPT-4o LLM. We generated vector embeddings from the 2015 American Thyroid Association Management Guidelines for Thyroid Nodules and Differentiated Thyroid Cancer and used vector similarity to determine which components of the guidelines would generate the most useful context for the introductory prompt based on the patient scenario. We also provided feedback generated from a pretest of 25 cases. After running the first assistant five times, we provided all five responses to a compounding assistant which took the most commonly appearing components of each and composited them together. We then evaluated the similarity between the expert A&P and the predicted A&P across the domains of (1) diagnosis, (2) the need for an operation, and (3) type of operation.

To evaluate EndoGPT, we measured concordance between the expert-generated and the predicted A&Ps across three domains: (1) diagnosis, (2) need for an operation, and (3) type of operation (Figure 1). This study was deemed exempt by the Columbia University Institutional Review Board (`Protocol AAAV1151`). Our code is available on GitHub.

## Results

We tested EndoGPT on 50 patient scenarios and achieved an overall concordance of 83%. EndoGPT agreed with the expert’s diagnosis completely in 44/50 cases and partially in 5/50 cases (93% concordant). Moreover, the assistant agreed with the expert’s need for an operation in 41/50 cases (82% concordant). When the expert recommended surgery (n=36 cases), the assistant agreed with the expert’s choice of operation completely in 24 cases and partially in two cases (69% concordant) (Figure 2). Details on the differences in A&Ps are described in Table S1.

**Figure 2:**
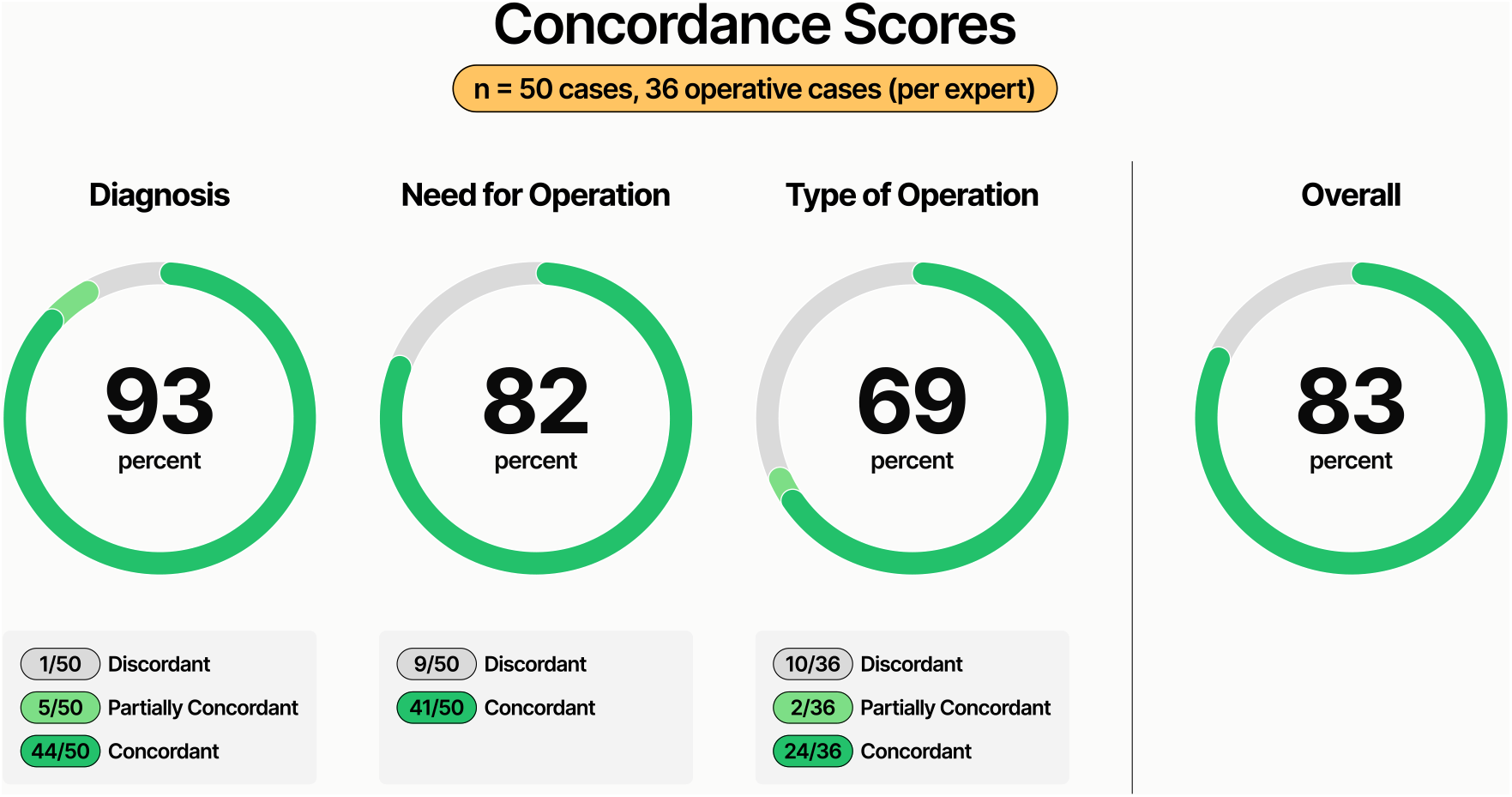
EndoGPT concordance scores in the domains of diagnosis, need for an operation, type of operation, and overall. When assessing concordance in diagnosis and operation type, we allowed partial credit for partially concordant responses.

## Discussion

Our early experience with EndoGPT suggests that surgeons who may not have the technical resources to build their own LLMs can still use general-purpose models like GPT-4o to develop clinical decision support tools. We achieved an 83% concordance with expert A&Ps using knowledge-retrieval and prompt engineering.

Our model was most concordant when predicting a diagnosis and least concordant when suggesting a specific operation. Specific areas of recurring discordance were in the type of lymph node dissection (LND) recommended (e.g. EndoGPT did not assign a laterality to central LND) or the recommendation of surgery for benign nodules causing compressive symptoms (rather than performing fine needle aspiration). The latter may have occurred because we gave EndoGPT specific feedback during pretesting to consider surgery for benign, compressive nodules, highlighting the risk of over-prompting the model. In some cases, because we tested concordance with a singular A&P, it is possible that EndoGPT suggested a safe alternative approach. Thus, we may be underestimating EndoGPT’s overall accuracy. In future experiments, a panel of experts can assess EndoGPT’s responses for accuracy.

Though not intended to replace physician evaluation, tools like EndoGPT may help train 4 surgical residents, assist non-specialist providers with initial workup and management, or make technical documents such as guidelines more accessible to patients. Utility will likely be greatest in areas of medicine where clear guidelines already exist. Further studies will be needed to fully optimize this system for patient care.

## Data Availability

Our data and code are available on GitHub.

https://github.com/tsathe/endogpt

## Supplementary Tables

**Table S1:** EndoGPT concordance scores in the domains of diagnosis (Dx), need for an operation (Op?), and type of operation (Type). When EndoGPT achieved a less than perfect score, we explain the areas of discordance. FNA = fine needle aspiration; PTC = papillary thyroid carcinoma; LND = lymph node dissection.

| Case | Dx | Op? | Type | Explanation of Discordance |
| --- | --- | --- | --- | --- |
| 1 | 2 | 1 | 2 |  |
| 2 | 2 | 1 | 2 |  |
| 3 | 2 | 1 |  |  |
| 4 | 1 | 1 | 2 | EndoGPT's diagnosis was a benign nodule, while the expert's diagnosis was a suspicious enlarging nodule for a nodule with Bethesda III classification on FNA. |
| 5 | 2 | 1 |  |  |
| 6 | 2 | 1 | 2 |  |
| 7 | 2 | 1 | 1 | EndoGPT recommended total thyroidectomy with a right lateral LND, while the expert recommended total thyroidectomy with a right central and lateral LND for PTC with a positive node. |
| 8 | 2 | 1 | 2 |  |
| 9 | 2 | 1 | 1 | EndoGPT recommended neck dissection, while the expert recommended neck dissection based on intraoperative findings for PTC with a suspicious node on imaging. |
| 10 | 2 | 1 | 2 |  |
| 11 | 1 | 0 |  | EndoGPT's diagnosis was a benign nodule and recommended lobectomy, while the expert's diagnosis was a nontoxic multinodular goiter and recommended FNA. |
| 12 | 2 | 0 |  | EndoGPT recommended completion lobectomy, while the expert recommended FNA for a multinodular goiter. |
| 13 | 2 | 1 | 2 |  |
| 14 | 2 | 1 | 2 |  |
| 15 | 2 | 0 |  | EndoGPT recommended lobectomy, while the expert recommended surveillance for a Bethesda III nodule with negative molecular testing. |
| 16 | 2 | 1 | 0 | EndoGPT recommended total thyroidectomy, while the expert recommended lobectomy for PTC in two ipsilateral nodules. |
| 17 | 2 | 1 | 2 |  |
| 18 | 2 | 1 | 2 |  |
| 19 | 2 | 1 | 0 | EndoGPT recommended lobectomy, while the expert recommended total thyroidectomy for a nodule in the setting of Graves disease. |
| 20 | 2 | 0 | 0 | EndoGPT recommended FNA, while the expert recommended total thyroidectomy for a multinodular goiter with growing nodules. |
| 21 | 2 | 1 | 0 | EndoGPT recommended lobectomy, while the expert recommended total thyroidectomy for a multinodular goiter. |
| 22 | 2 | 1 |  |  |
| 23 | 2 | 1 |  |  |
| 24 | 2 | 0 |  | EndoGPT recommended lobectomy, while the expert recommended repeat FNA for a growing nodule. |
| 25 | 2 | 1 | 2 |  |
| 26 | 2 | 1 | 2 |  |
| 27 | 2 | 1 | 0 | EndoGPT recommended total thyroidectomy, while the expert recommended lobectomy for a multinodular goiter. |
| 28 | 2 | 1 | 0 | EndoGPT recommended total thyroidectomy and central neck dissection, while the expert recommended lobectomy for a small PTC with a negative node on FNA. |
| 29 | 2 | 1 |  |  |
| 30 | 2 | 1 | 2 |  |
| 31 | 0 | 0 |  | EndoGPT's diagnosis was a goiter and recommended total thyroidectomy, while the expert's diagnosis was a nodule and recommended FNA. |
| 32 | 2 | 1 | 2 |  |
| 33 | 2 | 1 | 2 |  |
| 34 | 2 | 1 | 2 |  |
| 35 | 2 | 1 | 2 |  |
| 36 | 2 | 0 |  | EndoGPT recommended total thyroidectomy, while the expert recommended FNA for a growing nodule with compressive symptoms. |
| 37 | 2 | 1 |  |  |
| 38 | 2 | 1 | 2 |  |
| 39 | 2 | 1 | 2 |  |
| 40 | 2 | 0 |  | EndoGPT recommended lobectomy for a multinodular goiter, while the expert recommended FNA. |
| 41 | 1 | 1 | 0 | EndoGPT's diagnosis was a toxic multinodular goiter and recommended total thyroidectomy, while the expert's diagnosis was a toxic adenoma and recommended lobectomy. |
| 42 | 2 | 1 | 0 | EndoGPT recommended total thyroidectomy, while the expert recommended lobectomy for a multinodular goiter. |
| 43 | 1 | 1 | 0 | EndoGPT's diagnosis was PTC and recommended total thyroidectomy, while the expert's diagnosis was a suspicious nodule and recommended FNA. |
| 44 | 2 | 0 |  | EndoGPT recommended lobectomy, while the expert recommended repeat FNA for a Bethesda IV nodule. |
| 45 | 1 | 1 | 2 | EndoGPT's diagnosis was metastatic PTC, while the expert's diagnosis was PTC. |
| 46 | 2 | 1 | 2 |  |
| 47 | 2 | 1 | 2 |  |
| 48 | 2 | 1 | 2 |  |
| 49 | 2 | 1 | 0 | EndoGPT recommended completion left lateral neck dissection in a patient who previously had an excision of a mass found to be metastatic PTC, while the expert recommended total thyroidectomy and bilateral central and completion left lateral neck dissections. |
| 50 | 2 | 1 | 2 |  |

## References

[1] Harsha Nori, Yin Tat Lee, Sheng Zhang, Dean Carignan, Richard Edgar, Nicolo Fusi, Nicholas King, Jonathan Larson, Yuanzhi Li, Weishung Liu, Renqian Luo, Scott Mayer McKinney, Robert Osazuwa Ness, Hoifung Poon, Tao Qin, Naoto Usuyama, Chris White, and Eric Horvitz. Can generalist foundation models outcompete Special-Purpose tuning? case study in medicine. November 2023. URL http://arxiv.org/abs/2311.16452.

[2] Tejas S Sathe, Joshua Roshal, Ariana Naaseh, Joseph C L’Huillier, Sergio M Navarro, and Caitlin Silvestri. How I GPT it: Development of custom artificial intelligence (AI) chatbots for surgical education. J. Surg. Educ., 81(6):772–775, June 2024. ISSN 1931-7204, 1878-7452. doi: 10.1016/j.jsurg.2024.03.004. URL http://dx.doi.org/10.1016/j.jsurg.2024.03.004.

[3] Bryan R Haugen, Erik K Alexander, Keith C Bible, Gerard M Doherty, Susan J Mandel, Yuri E Nikiforov, Furio Pacini, Gregory W Randolph, Anna M Sawka, Martin Schlumberger, Kathryn G Schuff, Steven I Sherman, Julie Ann Sosa, David L Steward, R Michael Tuttle, and Leonard Wartofsky. 2015 american thyroid association management guidelines for adult patients with thyroid nodules and differentiated thyroid cancer: The american thyroid association guidelines task force on thyroid nodules and differentiated thyroid cancer. Thyroid, 26(1):1–133, January 2016. ISSN 1050-7256, 1557-9077. doi: 10.1089/thy.2015.0020. URL http://dx.doi.org/10.1089/thy.2015.0020.

[4] Underfitted. Building a RAG application from scratch using python, LangChain, and the OpenAI API, March 2024. URL https://www.youtube.com/watch?v=BrsocJb-fAo.

